# COMPARISON OF ARTIFICIAL INTELLIGENCE ENABLED METHODS IN THE COMPUTED TOMOGRAPHIC ASSESSMENT OF COVID-19 DISEASE

**DOI:** 10.1101/2020.09.02.20186650

**Authors:** Atul Kapoor, Goldaa Mahajan, Aprajita Kapoor

## Abstract

**Objectives:** Comparison of three different Artificial intelligence (AI) methods of assessment for patients undergoing Computed tomography (CT) for suspected Covid-19 disease. Parameters studied were probability of diagnosis, quantification of disease severity and the time to reach the diagnosis.

**Methods:** 107 consecutive patients of suspected Covid-19 patients were evaluated using the three AI methods labeled as AI-I,II, III alongwith visual analysis labeled as VT for predicting probability of Covid-19, determining CT severity score (CTSS) and index (CTSI), percentage opacification (PO) and high opacification (POHO). Sensitivity, specificity along with area under curves were estimated for each method and the CTSS and CTSI correlated using Friedman test.

**Results:** Out of 107 patients 71 patients were Covid-19 positive and 20 negative by RT-PCR while 16 did not get RT-PCR done. AI-III method showed higher sensitivity and specificity of 93% and 88% respectively to predict probability of Covid 19. It had 2 false positive patients of interstitial lung disease. AI-II method had sensitivity and specificity of 66% and 83% respectively while visual (VT) analysis showed sensitivity and specificity of 59.7% and 62% respectively. Statistically significant differences were also seen in CTSI and PO estimation between AI-I and III methods (p< 0.0001) with AI-III showing fastest time to calculate results.

**Conclusions:** AI-III method gave better results to make an accurate and quick diagnosis of the Covid-19 with AUC of 0.85 to predict probability of Covid-19 alongwith quantification of Covid-19 lesions in the form of PO, POHO as compared to other AI methods and also by visual analysis.

**KEY POINTS:** CT examinations of the chest can be more accurate and informative in detecting Covid-19 if combined with AI methods which are being designed to achieve this objective. In this study we compared three AI methods with Visual analysis and the results show.

- Al-III method had a higher sensitivity and specificity of 93% and 88% compared to other methods in predicting probability of Covid-19.
- Significant inter method variations were seen in quantifying Covid-19 opacities as CTSS,CTSI, PO and POHO variables (p< 0.0001). AI-III method showed no statistical difference with VT method for PO variable (p = 0.24) and was the only method which depicted all the variables..
- Time to processing results was the shortest with AI-III method.

## INTRODUCTION

Covid-19 is a highly infectious disease affecting more than 3 million people worldwide as on this date. Due to its high rate of infectiousness rapid tools are needed for early and accurate diagnosis and also for monitoring the progress of disease (1). So far RT-PCR test using throat and nasal swabs form the backbone of diagnosis and is considered to be gold standard but is marred by reduced sensitivity. It has high false negative rates i.e.30–35%, is more time consuming and also has false positives(2). The epidemic is thus driving researchers to think of more efficient ways to cope with the huge demand for diagnosis. Imaging has been used in China on a large scale to tackle the endemic and there have been numerous reports about the experiences using CT scans. Chinese national guidelines have recommended CT scan as a key method to diagnose Covid-19(3). Typical reported features on CT of Covid-19 include multifocal ground glass opacities and consolidations with peripheral and basal predilection(4,5).Based on these Radiological society of North America has proposed CORADS classification system on a scale of 1–6(6,7). However American college of Radiology in June 2020 issued guidelines mitigating its use for diagnosis of COVID-19 mainly due to fear of contaminating radiology facilities and also due to its lack of specificity(8). So what should be the role of radiology in current pandemic is a question under debate (9,10). Many researchers have come up with Artificial intelligence(AI) based prototypes to automate the diagnosis of Covid-19 disease which will expedite early and accurate diagnosis. This should help physicians triage patients into Covid-19 designated units for treatment and also help contain further spread of disease(11).

We evaluated the results of three such AI based methods using Computed tomography images of the chest done on patients suspected to be having Covid-19 disease with following objectives

a. To estimate the ability of AI methods to diagnose Covid-19 disease. b) Compare the results of different AI methods and with visual analysis in determining the severity of disease. c) To determine the time taken for making diagnosis using AI.

## MATERIAL AND METHODS

AI based methods using computed tomographic images of 107 consecutive patients suspected of having Covid-19 disease were included in the study after obtaining consent from local ethics review committee and informed consent from the patients. Plain computed tomographic examination was done on 128 slice Siemens Healthineers Go top system using standard operating parameters with tube voltage of 80KV, 240ma, 1.5 mm slice thickness and reconstruction kernel of B40,60. Medical records of all patients were also examined alongwith chief complaints and available laboratory data and presence of any comorbid disease was recorded. Short follow up was done in all patients for knowing the RT- PCR and clinical status of all patients from their families and treating physicians.All necessary precautions regarding using of personal protective equipment, disinfection of the scan room and department were also taken before and after each examination and patients referred to respective medical units.

The imaging data was then transferred to local PACS and then sent to the collaborators for analysis. Collaborations were done with two AI software companies to process the data. A) AI-I label was given to AI method using COVID-19 AI software from Thirona. B) AI-II label was for method from Quibim,. These were cloud based methods with analysis being done servers of the respective vendors. C) AI-II label was for onsite Pneumonia analysis from Siemens Healthineers done on Siemens Syngo Via system. D) VT (Visual truth) label consisted of visual analysis by two radiologists on PACS and conclusions were based by consensus.

The following parameters were evaluated for each patient using all four methods. A) Diagnostic probability of Covid-19 disease. B) Grading the severity of disease in terms of CT severity scale(CTSS) and CT severity scale index(CTSSI), percentage opacification (PO), percentage high opacification (POHO) and dominant lobe involvement (Figures1A-D).C)Mean time taken for each evaluation by all four methods on per patient basis was recorded. This included time taken from loading the data to completion of results on per patient basis by all methods. The CTSS score of degree of lung involvement was calculated as follows based on method by Bernheim(5). AI –I method used a score of 0–25 based on a scale of 0–5 as follows • 0: lobe is not affected. • 1: 1–19% of the lobe is affected. • 2: 20–39% of the lobe is affected. • 3: 40–59% of the lobe is affected. • 4: 60–79% of the lobe is affected 5.80–100%. In AI-III CTSS score was 0–20 as follows • 0: lobe is not affected. • 1: 1–25% of the lobe is affected. • 2: 25–50% of the lobe is affected. • 3: 50–75% of the lobe is affected. • 4: 75–100% of the lobe is affected. In AI-I method no severity scoring was done. VT method used a score of 0–20 based on method of dividing lungs into superoinferior and mediolateral quadrants on coronal views A score of 0–20 was assigned on a scale of 0–5 for each quadrant based on percentage of lung involved from 0–20%, 20–40%,40–60%,60–80% and 80–100% opacities respectively. CTSSI was calculated for each score to compare the scores determined by each method as:

CTSSI: CTSS/Total Score x100%.

a. Percentage of opacity (POO) was calculated as follows:

PO = 100× Volume of predicted abnormalities of lung / volume of lung mask.
b. Percentage of high opacity (POHO): 100xvolume of predicted high opacity/volume of lung mask.
c. Probability of Covid-19 was depicted based on Area under curve method and a threshold value of more than 0.85 was deemed as having Covid-19 based on AI-II,III methods while VT method probability was based on visual analysis and expertise by both consensus readers.
d. Pattern of distribution of lung lesions was determined as lobar, unilateral or bilateral or whole lung involvement.

**Figure 1:**
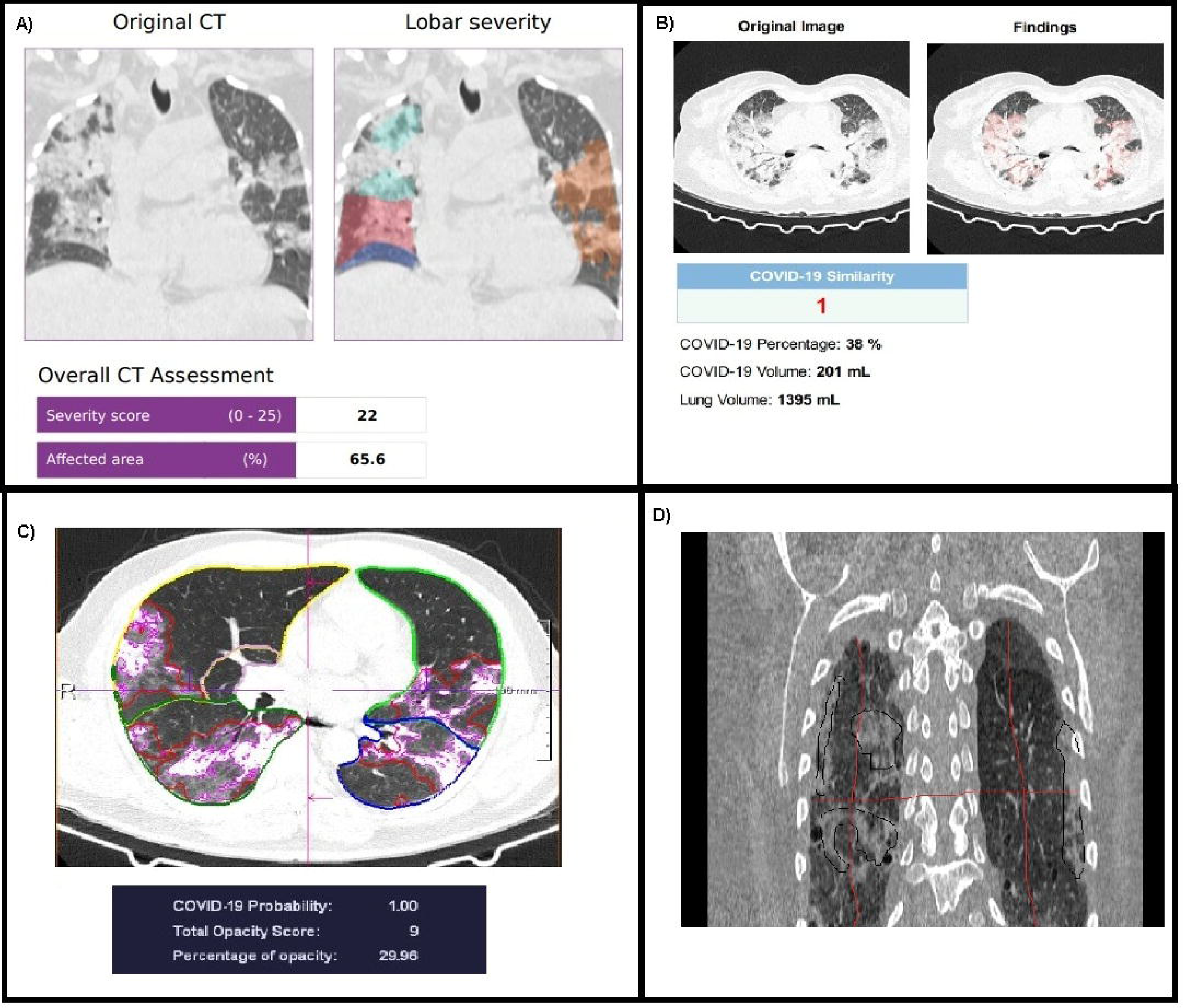
A): Plain CT evaluation by AI-I method showing CTSS of 22 and CTSI 88% with percentage opacification of 65.6%.B) AI-II showing indeterminate CTSS and CTSI with PO of 38% and Covid probability of 1.0. C) AI-III in same patient with CTSS and CTSI 9 and 45% with Covid probability of 1.0 with PO of 38% d) VT analysis image with CTSS and CTSI of 8 and 40%.

All the results were statistically analysed using Analyse-IT software (Leeds, UK) to determine the parametric of mean, standard deviation, correlation was done using Friedman test for pair groups between all four methods and Sensitivity, specificity along with Area under curves calculated.

## RESULTS

The study comprised of consecutive 107 patients of suspected Covid-19 whose computed tomography examinations showed focal or diffuse ground glass opacities and had clinical presentations suggestive of Covid-19 disease. The demographic data of patients is listed in (Table 1). The mean age of patients was 53.6 years (51.1–56.1, 95% CI). Out of these 72 were males 35 females. 23 patients had co morbidities of which diabetes mellitus was the most common. RT-PCR was done in 91 patients out of which 71 were positive, 20 negative and in 16 patients it could not be done.

1. Probability of Covid-19 disease.

In the AI- I method there was no algorithm to calculate Covid-19 probability.
The AI-II method calculated Covid-19 probability with sensitivity and specificity of 63% and 83% with high false negative of 37% and false positive of 16.7% with a positive predictive value of 93%and negative predictive value of 35.7% (Figures 2A-C). Data of 18 patients in the programme could not be processed and was labeled as negative probability of 0.
AI-III method: Showed a sensitivity and specificity of 93% and 88% respectively with false positive and negative of 11% and 6% with a high positive predictive value of 97.2% and negative predictive value of 75%(Figures3 A-C). There were only 2 false positive cases all of whom had prior interstitial lung disease. 5 false negative cases were seen with normal CT.
VT method showed a sensitivity and specificity of 59.7% and 62% to diagnose Covid-19 with false positive and false negative of 37% and 40% respectively with positive predictive value of 83% and negative predictive value of 37%(table2). The area under curve estimation showed for AI-II,III and VT methods as 0.68, 0.85and 0.62 respectively with differences between AI-III and AI-II and VT method being statistically significant(p-0.0001).(Figure4).
2. CTSS and CTSI estimation:

AI,III methods could calculate CTSS and CTSI. AI-I method determined the CTSS in all patients with median CTSS of 12 (10–13 96.7%) and CTSI of 48% (median 40–52% 96.7% CI). AI-III method and VT methods also determined the CTSS and CTSI with mean scores of 7 (6–9 96.7% CI), 35% (30–45 97.6% CI)for AI-III method and 6.50(6–9 96.7%CI) 32.5 (30–45 96.7%CI) respectively. Friedman test was done between three methods of analysis and showed statistically significant differences between AI-III method and AI-I and VT methodsp< 0.0001(Table3),(Figures 5A-D).
3. Time estimation for results: Since AI-I,II methods were cloud based methods the average processing time was 10 minutes, 25 minutes respectively with AI-II showing maximum number of incomplete processing results i.e. 18 patients. AI-III method showed the quickest processing time of 2 minutes and had the advantage of being onsite method.
4. Percentage opacities(Figure5): AI-III method showed median percentage opacification of 21.0 (median 15–30 96–7%CI) while AI-II underestimated percentage opacities with median value of 1.0 while AI-I and VT method did not dlineate percent opacification.
5. Percentage of high opacities: was estimated only estimated by AI-III method with a median of 4.0 (2.5–5.2 96.7%CI).
6. Lobar dominance was computed by all the methods with commonest patterns being bilateral lower lobe predominance followed by complete lung involvement.

**Table 1:** Showing patient demographics.

| TABLE 1: |  | Patient demographics |  |
| --- | --- | --- | --- |
| Sno | Parameters | Median | CI |
| 1 | Age(years) | 55 | 53-57 |
| 2 | Sex |  |  |
|  | males | 72 |  |
|  | females | 35 |  |
| 3 | COMORBIDITIES | 28 |  |
| 4 | Chief Complaints |  |  |
|  | fever | 85 |  |
|  | Breathlessness | 70 |  |
|  | Dry cough | 55 |  |
|  | Pain abdomen | 14 |  |
|  | Diarrhoea | 19 |  |
|  | Loss of smell | 8 |  |
|  | Body aches | 80 |  |
| 5 | Spo2 mm HG | 94 |  |
| 6 | Prior Investigations |  |  |
|  | Xray Chest | 90 |  |
|  | Hemogram | 65 |  |
|  | CRP | 24 |  |

**Figure 2:**
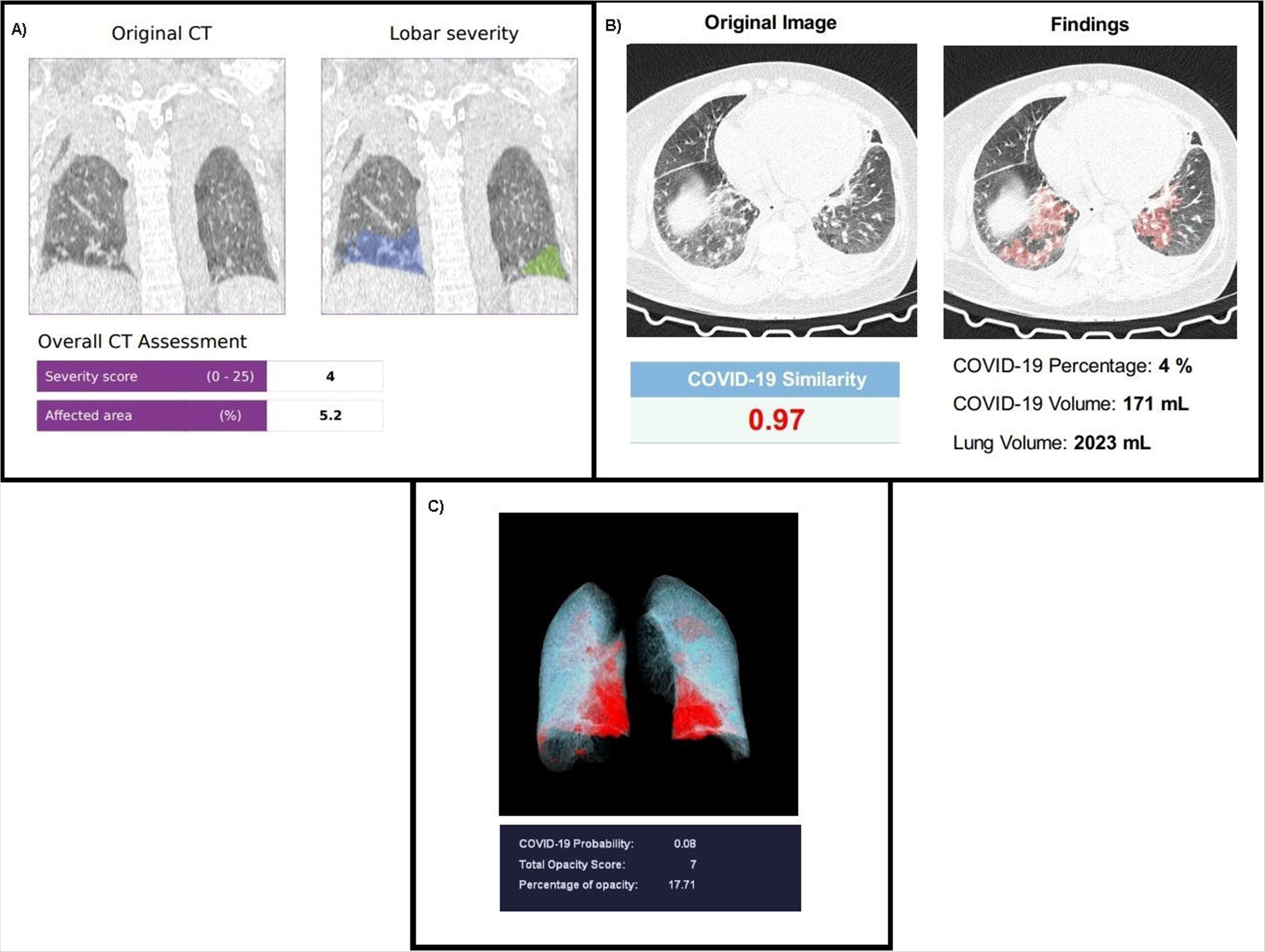
A): Plain CT image of non covid patient with pulmonary edema evaluated by Al-I showing CTSI of 4, CTSI 16% with PO of 5.2%. B) Al-II showing false positive Covid probability of 0.97 with PO of 4%. C) Al-III showing paracardiac central opacities with Covid probability of 0.08, CTSI of 7, CTSI 35% and PO 17%

**Figure 3:**
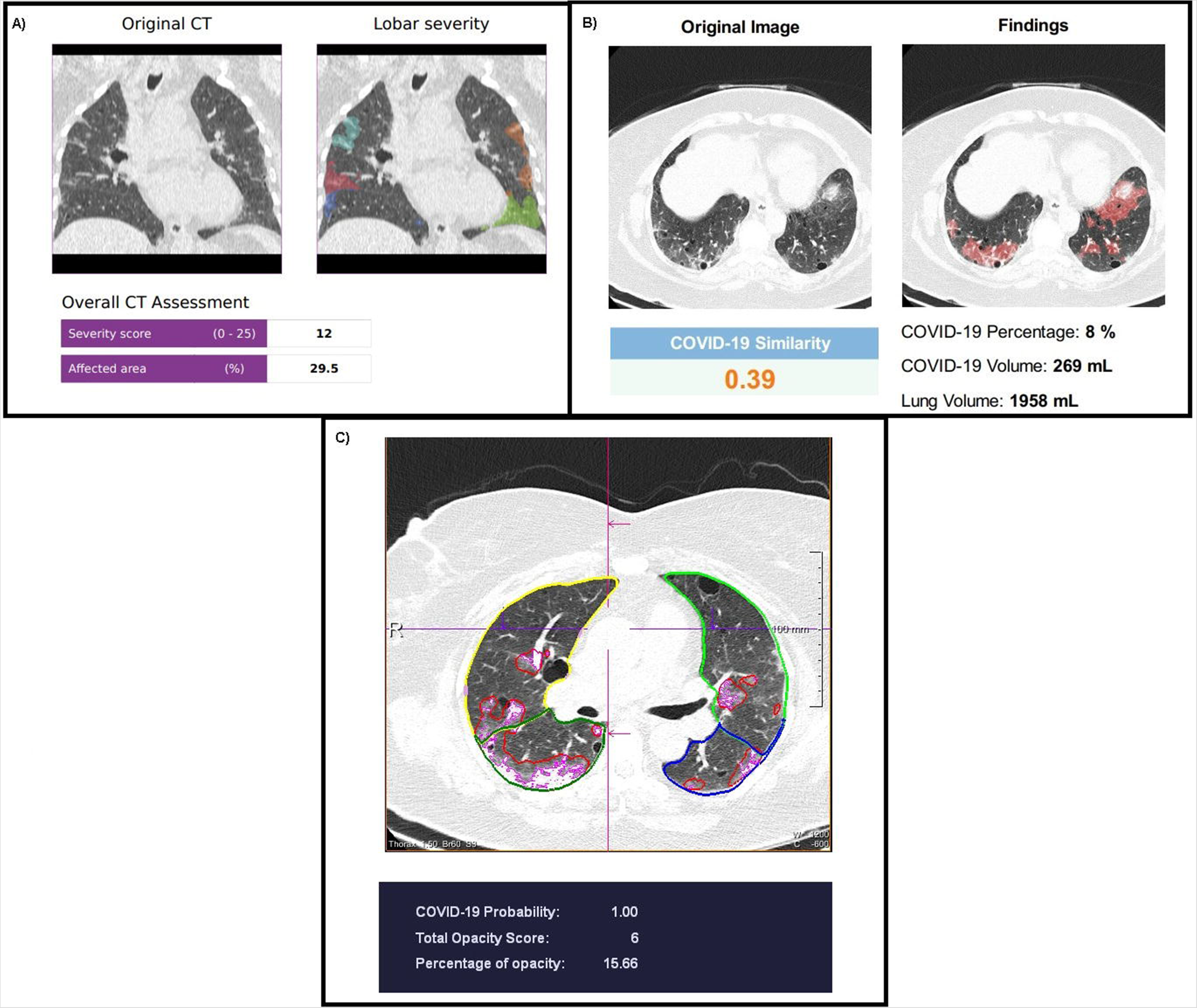
A): RT-PCR positive patient evaluated on Al-I showing CTSS of 12,CTSI 48% with percentage opacification of 29.5% on Al-I B) Al-II method in same patient showing PO 8% with false negative Covid probabillity. of 0.39,.C) Al-III shows CTSS6, CTSI 30% with Covid probability of 1.0 WITH PO 15.6%

**Table 2:**
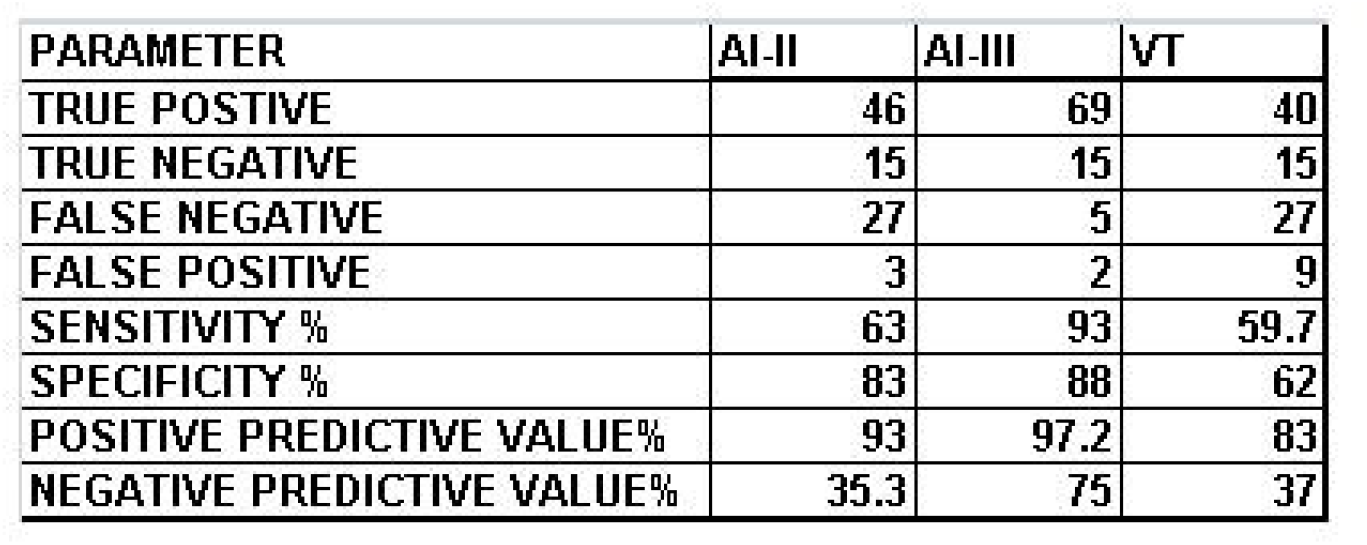
Showing qualitative parameters of diagnostic evaluation of Al-I,III and VT methods.

| PARAMETER | AI-II | AI-III | VT |
| --- | --- | --- | --- |
| TRUE POSTIVE | 46 | 69 | 40 |
| TRUE NEGATIVE | 15 | 15 | 15 |
| FALSE NEGATIVE | 27 | 5 | 27 |
| FALSE POSITIVE | 3 | 2 | 9 |
| SENSITIVITY % | 63 | 93 | 59.7 |
| SPECIFICITY % | 83 | 88 | 62 |
| POSITIVE PREDICTIVE VALUE% | 93 | 97.2 | 83 |
| NEGATIVE PREDICTIVE VALUE% | 35.3 | 75 | 37 |

**Figure 4:**
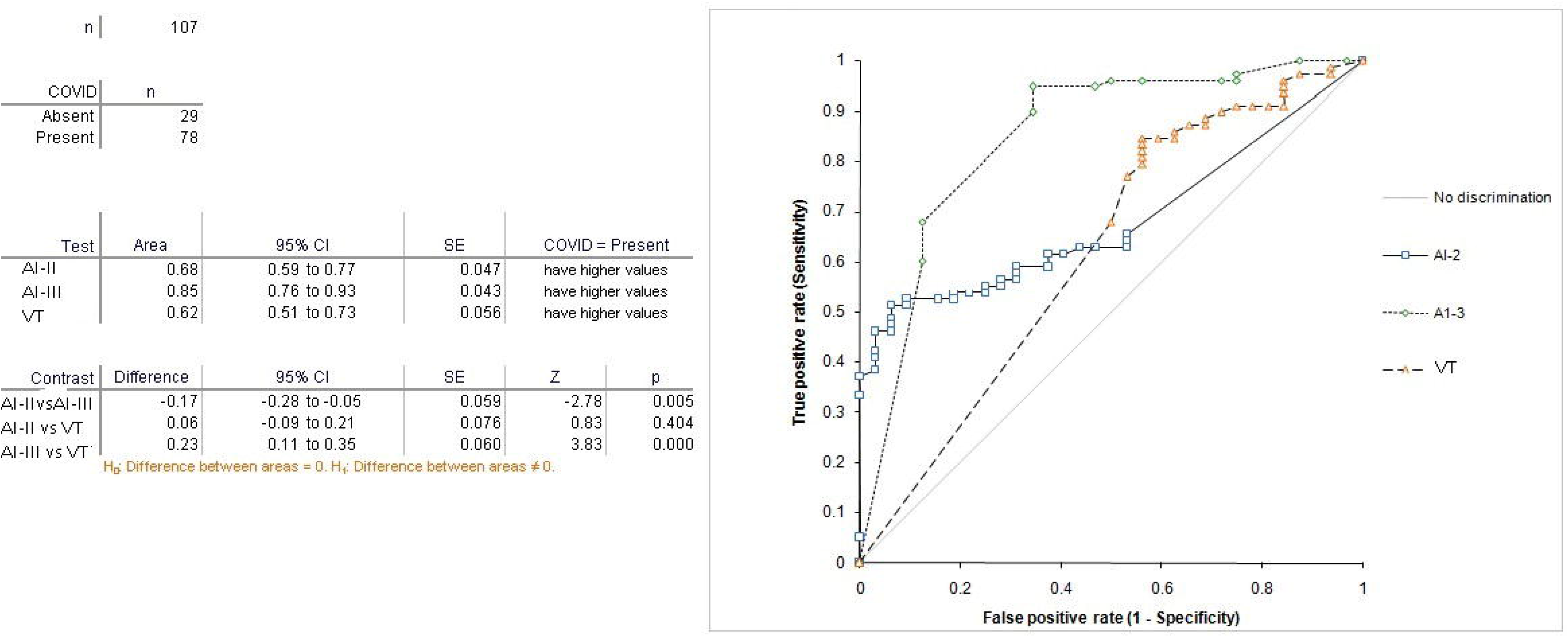
Showing AUC,s for all the three AI methods and VT method

**Table 3:** Friedman test of CTSI of Al-I,III and VT method.

n | 107
|  | n | Rank sum | Mean rank |
| --- | --- | --- | --- |
| AI1-CTSSI | 107 | 304.0 | 2.84 |
| AI-III CTSSI2 | 107 | 113.0 | 1.06 |
| AI-IV CTSSI | 107 | 225.0 | 2.10 |

Friedman's statistic | 178.42
$\chi^2$ statistic | 178.42
DF | 2
p | <0.0001 (chisqr approximation, corrected for ties)

**Figure 5:**
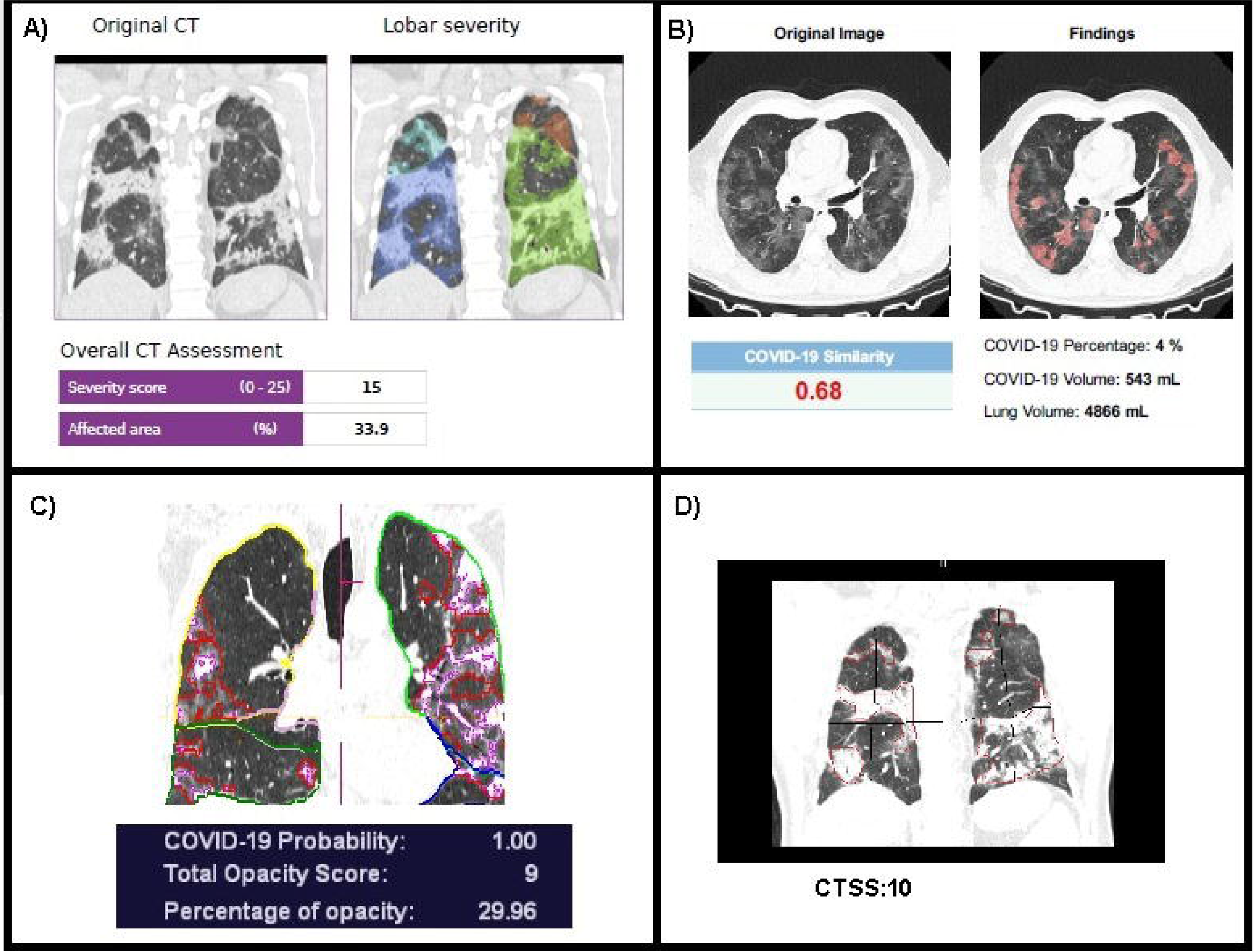
A): Al-I showing a RT PCR positive patient with PO of 33.9% and CTSS 15, CTSI 60%. B) A PO of 4% with Covid similarity of 0.68 seen by Al-II. C) Al-III shows PO of 29% with CTSS and CTSI of 9 and 45% with Covid probability 1.0. D) VT method correlates CTSS 10 and CTSI of 50%.

RT-PCR was done in 91 patients while 16 patients did not get RT-PCR, out of these there were 42 positive results for Covid-19 detection in the first test and 29 in second test while 20 were repeatedly negative. The sensitivity and specificity of AI- methods and VT method were calculated based on the 91 results of RT-PCR.

## DISCUSSION

CT imaging has a important undeniable role in the evaluation of patients with lower respiratory tract disease including suspected COVID-19. Evaluation of a suspected Covid-19 patient can be done by chest CT to answer several questions in a clinical setting i.e, from diagnosis, to determining the disease severity, progression and treatment response. In a pandemic situation timely diagnosis by the use of imaging can be the essence of management(12). Our study shows that use of AI with CT does improve the sensitivity and specificity of diagnosis compared with visual analysis method which was the primary objective of the study. It is critically important to have probability algorithm in the AI methods to be able to answer the question of whether Covid-19 or not. Our study shows that at this point of time not all AI methods have this ability which was absent with in AI- I method and variations in predicting the probability were also seen in the results between AI-II, III methods. AI-III showed the highest sensitivity and specificity of 93% and 88% respectively suggesting it is almost ready for clinical use with results being superior to visual analysis alone. The probable reason for this is the machine learning method used in this technique. The developers of this AI method used a supervised deep learning method based on 3D neural network in which a two channel 3D tensor was used where the first channel contained CT Hounsfield units masked by lung segmentation while the second channel had probability maps of opacity classifiers of metric data (13,14). Chiganti etal (13) showed improved performance with AUC of 0.90(0.85–0.94 95%CI with sensitivity and specificity of 86% and 81% respectively which was similar to the results seen in the present study. The earlier AI methods used metric based AI classifiers based on opacity metrics with generation of heat maps and regression algorithms with reduced sensitivity specificity of 74% as was with AI-II method in this study. Similar results have also been shown by Li etal (15) who showed reduced false positive and negatives in making diagnosis with the use of 3D machine learning methods. CTSS has recently been proposed as a reliable measure to demonstrate correlation of the severity of disease with clinical condition of patient (16). It has been used to assess the progression of disease by many studies (17). In the present study accurate CTSS scoring was possible with both AI-I and AI-III methods but only AI-III method shows no statistical difference with the VT method (p = 0.24). AI-II method could not compute a severity score. In our study lung and lobar and opacity segmentations were also possible with the use of all AI based methods. However AI-III method used deep image to image network machine learning algorithms which were more robust and depicted both PO and POHO estimates which was not possible by other AI methods nor by visual method. These quantitative analysis of the severity of lung involvement help physicians to triage and monitor Covid –19 patients and are essential in all AI methods of analysis (18,19).Findings of the present study suggest that standardized objective measures of disease evaluation were missing in some of the methods evaluated in this study therefore improved training data sets and algorithms are required in AI-I,II methods to achieve accurate segmentation before all AI based methods can be applied in routine clinical practice (20). This would mean that some more time is required before AI comes into clinical use. The third objective of the study i.e determining time to diagnosis was also achieved in the study with AI-III method which had a mean processing time of 2 minutes which was quick enough to make the diagnosis of Covid-19. To deal with highly infectious disease like Covid-19 early diagnosis is the essence and onsite processing algorithms like used in AI-III should be preferred than cloud based evaluation techniques.

To conclude addition of AI along with CT chest evaluation looks attractive and its use achieves the objectives set for evaluation in the present study especially with the use of AI-III method. It has the potential to improve the accuracy and time to make the diagnosis. The study also highlights limitations in various AI methods tested along with inter method variations of results like estimation of CTSI, percentage opacification and some more time may be required before it comes into clinical use. Out of the AI techniques compared AI-III method appears to be more advantageous and accurate compared with other AI methods including the visual method alone.

## Data Availability

institutional data server

https://www.advanceddiagnostics.in

## ABBREVIATIONS

PO: percentage opacification
POHO: percentage opacification of high opacities.
AI: Artificial intelligence
CTSS: CT severity score
CTSI: CT severity score index.
CT: Computed tomography.
VT: visual truth.

## Acknowledgments

*BLINDED*

